# Benchmarking HLA genotyping from whole genome sequencing across multiple sequencing technologies

**DOI:** 10.64898/2026.02.10.26345621

**Authors:** Conor Cremin, Santosh Elavalli, Luis F Paulin, Judith Arres, Abdelrahman Ahmed Yehia Abdelaziz Saad, Azza Attia, Cyla Minas, Fatmah Aldhuhoori, Gurunath Katagi, Haiguo Wu, Hatim Sidahmed, Joseph Mafofo, Omar Soliman, Shalini Behl, Sharika Pariyachery, Vidhi Gupta, Duaa Ghanem, Hanil Sajjad, Thyago Cardoso, Albarah El-Khani, Fahed Al Marzooqi, Tiago Magalhaes, Fritz J Sedlazeck, Javier Quilez

**Affiliations:** M42, Abu Dhabi, United Arab Emirates; Baylor College of Medicine, Houston, TX, USA; Department of Molecular and Human Genetics, Baylor College of Medicine, TX, USA; Department of Computer Science, Rice University, 6100 Main Street, Houston, TX, USA

**Keywords:** HLA genotyping benchmarking, Short- and long-read WGS, Cross-platform performance evaluation, Bioinformatics tool assessment

## Abstract

**Background:** The hyper⍰polymorphic nature and structural complexity of the human leukocyte antigen (HLA) genomic region present challenges for accurate and scalable typing across diverse sample types. While whole⍰genome sequencing (WGS) offers the opportunity to infer HLA genotypes without targeted enrichment, systematic benchmarks across sequencing platforms, biospecimens and coverage levels remain limited.

**Results:** We assembled a multi-platform resource of WGS datasets derived from short-read (Illumina, MGI) and long-read (Oxford Nanopore Technologies R9 and R10) sequencing, spanning 29 biospecimens including cell lines, blood, buccal swab and saliva. We evaluated the performance of the HLA caller HLA*LA across 13 HLA genes, using a clinically validated assay as reference. WGS⍰based HLA genotyping achieved ∼95% accuracy across sequencing platforms, with Class I loci exhibiting higher accuracy than Class II. Cross⍰platform concordance was high, and performance remained consistent across Illumina, MGI and Oxford Nanopore chemistries. Analysis of blood, buccal swab and saliva samples showed that blood and buccal swabs supported accurate HLA inference, whereas saliva yielded reduced concordance. Down⍰sampling experiments demonstrated that 15x coverage was sufficient to retain >95% accuracy at two⍰field resolution, with lower depths supporting lower-resolution typing.

**Conclusions:** Our results demonstrate that WGS provides a robust, platform⍰agnostic framework for accurate HLA genotyping across sample types and coverage levels. These benchmarks establish practical conditions for reliable HLA inference and underscore the utility of WGS for population⍰scale HLA analyses and future clinical applications.

## Background

The human leukocyte antigen (HLA) locus within the major histocompatibility complex (MHC) encodes molecules that present peptides to T cells and underlies key immunological processes including transplant compatibility, infection control, autoimmunity and cancer immunity (1,2). Spanning ∼3-5 megabases (Mb) on chromosome 6p21.3, the HLA locus is one of the most polymorphic and structurally complex regions of the human genome, characterized by dense sequence homology, extensive paralogy, and complex linkage disequilibrium that complicate accurate genotyping (3). This complexity has historically limited comprehensive characterization, particularly in large-scale or clinical genomics settings.

Early HLA typing relied on serological assays followed by molecular typing based on polymerase chain reaction (PCR) and later Sanger sequencing (4). While resolution improved over time, these methods remained limited in scalability and genome coverage. High-throughput sequencing (HTS) increased throughput and resolution, with targeted HLA typing solutions existing for both short- and long-read sequencing technologies (5–8). However, most assays predominantly rely on locus-specific amplification and therefore remain susceptible to incomplete reference coverage, primer bias and allele dropout (9,10).

As population and clinical genomics scale up, large cohorts increasingly couple rich phenotypes to WGS, creating an opportunity to infer high⍰resolution HLA types directly from WGS rather than dedicated assays. In addition to enabling reutilization of existing genomics datasets, WGS captures all classical and non-classical HLA genes, including loci rarely interrogated by targeted approaches. As WGS becomes increasingly routine, these capabilities underscore the need for the broader genomics community, not only HLA specialists, to more systematically integrate HLA variation into phenotype⍰driven analyses.

However, accurate HLA inference from short reads remains challenging, especially for HLA Class II genes due to paralogous genes, long introns, and repetitive sequences (11). Recent algorithmic advances seek to overcome these challenges by aligning reads to variation graphs or population reference graphs (8,12). For instance, the HLA*LA caller introduced a linearly projected graph-alignment model that enables base-pair resolution across 13 major Class I and II genes (8). HLA*LA is among the tools that achieve high accuracy on both whole-exome and whole-genome sequencing data across diverse short- and long-read platforms, though multiple methods now support those data types (12–16). Yet independent benchmarks across sequencing technologies and depths as well as using diverse real-world sample types remain limited (17,18).

The goal of this study was to evaluate the utility of WGS across sequencing technologies to accurately genotype HLA loci. We first needed to confirm the performance of HLA*LA in an independent larger dataset than in the initial benchmark, which used only cell lines and an outdated long-read sequencing (LRS) technology. We then used well-characterized cell lines to evaluate the accuracy of our in-house end-to-end workflow for HLA typing from WGS for different sequencing platforms. We also evaluated the WGS-based HLA typing performance across platforms for real-world biobank samples (including blood but also less invasive buccal swabs and saliva). Finally, we assessed the sensitivity of HLA typing to sequencing coverage.

## Results

### A resource of 186 sequencing datasets for HLA profiling benchmarks

We built a panel of 29 biological specimens (24 Coriell cell lines and 5 human subjects) to generate HLA data across multiple DNA sources, sequencing platforms, and analysis tools (**Figure 1** and **Table 1**). For each of the 24 Coriell cell lines, we performed ∼30x WGS using three HTS: Illumina and MGI that are both short-read sequencing (SRS) and the long-read sequencing (LRS) technology Oxford Nanopore Technologies (ONT). ONT datasets included both R9 and higher-accuracy R10 chemistries. For the 5 human participants, we extracted DNA from three sample types (blood, buccal swab and saliva) and generated ∼30x WGS datasets with each of the three sequencing strategies (Illumina, MGI and ONT R9). All samples reached the target genome coverage (Average median = 33.4x; range = 11.0-50.0x).

**Figure 1.**
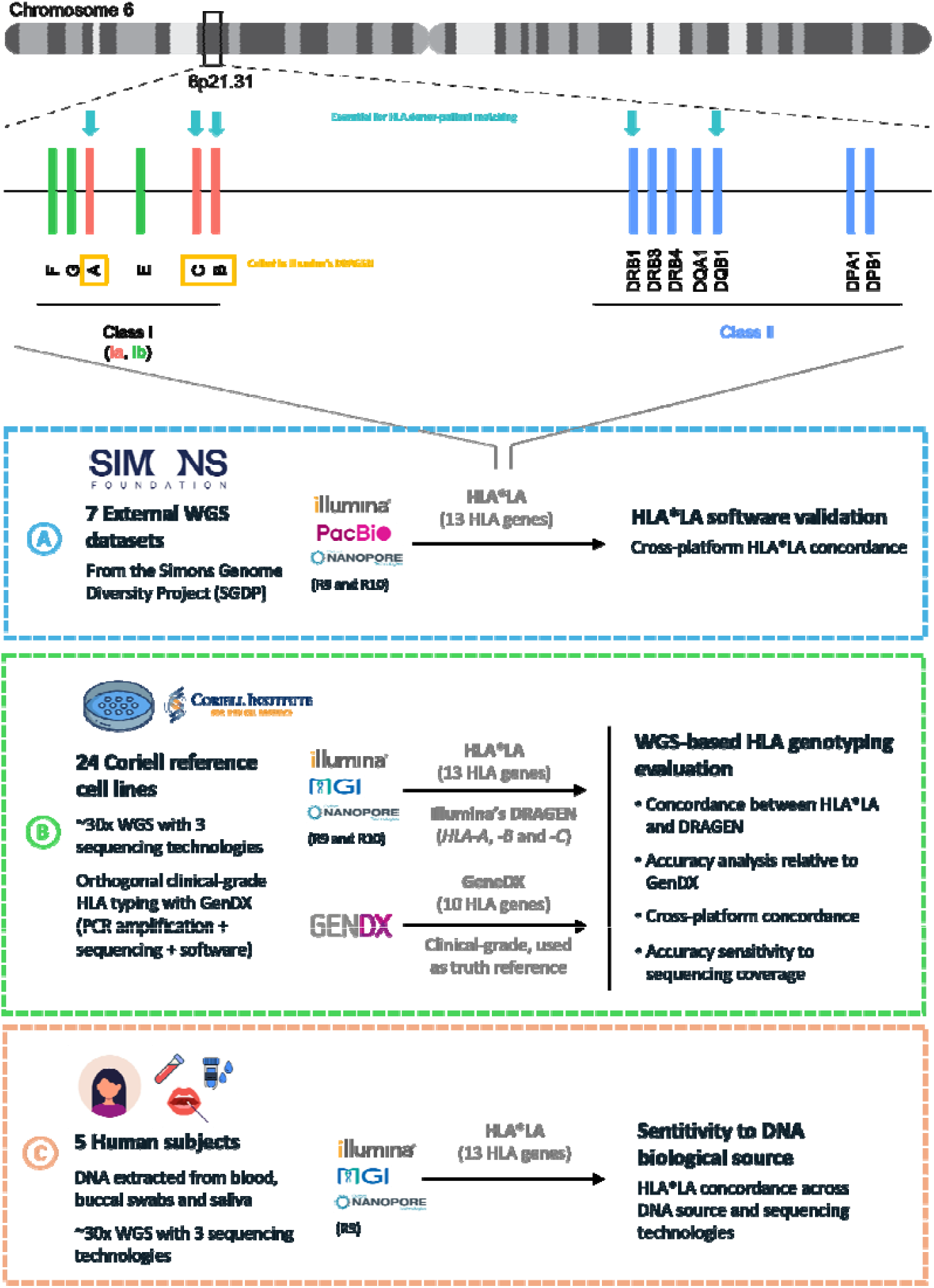
Overview of the WGS datasets and analyses. On the top, schematic representation of the HLA region on chromosome 6, indicating the 14 classical Class I and Class II HLA genes genotyped by HLA*LA. Also highlighted are genes covered by Illumina’s DRAGEN software (at the moment of the analysis) as well as those considered essential for HLA-based donor-patient matching.

**Table 1.** Summary of WGS and HLA coverage across datasets.

| Dataset | Sequencing Platform | Sample Type | WGS coverage |  |  | HLA coverage |  |  | Concordance |  |
| --- | --- | --- | --- | --- | --- | --- | --- | --- | --- | --- |
|  |  |  | Min | Median | Max | Min | Median | Max | Mean | SD |
| A. SDGP | Illumina | Cell line | N/A | N/A | N/A | 33.7 | 35.5 | 43.8 | 0.874 | 0.048 |
|  | PacBio | Cell line | ~30 | 39.7 | 50.0 | 43.1 | 45.5 | 51.1 | 0.879 | 0.052 |
|  | ONT (R9) | Cell line | ~30 | ~50 | ~60 | 39.1 | 52.0 | 68.6 | 0.879 | 0.052 |
| B. Coriell | Illumina | Cell line | 26.1 | 33.7 | 45.3 | 27.1 | 38.0 | 47.8 | 0.915 | 0.049 |
|  | MGI | Cell line | 35.3 | 36.7 | 58.4 | 25.9 | 31.1 | 48.9 | 0.915 | 0.049 |
|  | ONT (R9) | Cell line | 39.0 | 45.0 | 52.0 | 35.8 | 45.5 | 54.9 | 0.913 | 0.051 |
|  | ONT (R10) | Cell line | 23.0 | 31.0 | 65.0 | 23.9 | 33.5 | 69.3 | 0.913 | 0.051 |
| C. Cross samples | Illumina | Blood | 30.5 | 36.1 | 44.4 | 27.3 | 30.2 | 42.0 | 0.910 | 0.040 |
|  |  | Buccal swab | 25.6 | 27.1 | 39.6 | 24.1 | 28.9 | 36.0 | 0.910 | 0.040 |
|  |  | Saliva | 14.7 | 32.5 | 48.7 | 16.3 | 33.5 | 42.2 | 0.920 | 0.047 |
|  | MGI | Blood | 39.9 | 40.7 | 45.8 | 25.9 | 27.2 | 29.9 | 0.910 | 0.050 |
|  |  | Buccal swab | 28.7 | 41.3 | 46.4 | 31.4 | 38.5 | 40.9 | 0.920 | 0.047 |
|  |  | Saliva | 19.5 | 36.9 | 44.7 | 16.5 | 28.9 | 35.2 | 0.923 | 0.047 |
|  | ONT (R9) | Blood | 28.0 | 33.0 | 37.0 | 26.0 | 33.3 | 35.7 | 0.915 | 0.050 |
|  |  | Buccal swab | 20.0 | 23.0 | 26.0 | 21.5 | 23.7 | 25.9 | 0.923 | 0.054 |
|  |  | Saliva | 7.0 | 11.0 | 54.0 | 6.2 | 15.1 | 23.0 | 0.915 | 0.050 |

For each WGS dataset in **Table 1**, we inferred HLA genotypes across 13 major HLA genes using the HLA*LA software (8) and found high-confidence calls based on the HLA*LA quality scores (Q1: median = 1.00; range = 0.001 – 1.00). Additionally, we typed all Coriell cell lines with GenDX NGSengine, a clinically validated sequencing-based tool widely adopted for high-resolution HLA typing in both diagnostic and research settings (19,20); specifically, the GenDX assay uses PCR amplification followed by Illumina sequencing. In total, we generated 186 new datasets and incorporated publicly available WGS data from 7 samples from the Simons Genome Diversity Project (SGDP) sequenced with Illumina, ONT (R9 flow-cell) and Pacific Biosciences (PacBio) High Fidelity (HiFi) technologies. All HLA*LA allele calls are in **Supplementary Table 7**.

### Evaluation of WGS-based HLA genotyping

Our HLA typing analysis pipeline is based on the HLA*LA caller (**Figure 1**). Dilthey et al. previously demonstrated strong HLA*LA performance across sequencing technologies, but these benchmarks were largely limited to 1000 Genomes Project cell lines generated using sequencing technologies from over a decade ago (8). To evaluate whether HLA*LA remains robust on contemporary datasets and long-read chemistries, we first assessed cross-platform reproducibility using publicly available WGS data from seven individuals in the SGDP, sequenced on Illumina, Pacific Biosciences (PacBio) HiFi, and Oxford Nanopore Technologies (ONT) R9 platforms (**Figure 1A**).

HLA*LA maintained high concordance across Illumina, PacBio HiFi, and ONT WGS datasets (mean 93.9%), with half of the loci achieving 100% agreement and most others exceeding 78% (**Supplementary Results**). Notably, discrepancies were locus⍰specific rather than technology⍰driven and showed no association with coverage or caller confidence. Together, these results demonstrate that HLA*LA delivers highly reproducible, platform-agnostic HLA typing across both Class I and Class II loci from WGS data, supporting its use as the core caller in our downstream analyses.

We next expanded our evaluation by integrating HLA*LA into our in-house short- and long-read WGS workflows and benchmarking performance using the 24 Coriell cell lines. Using Illumina WGS data, we first compared HLA*LA against Illumina’s DRAGEN HLA caller, a widely used reference method for clinical-grade sequence analysis (7) (**Figure 1B**). Because DRAGEN supported only *HLA-A, -B*, and *-C* at the time of analysis, this comparison was restricted to Class I loci. Concordance was exceptionally high (mean pairwise concordance = 99.3%, **Supplementary Table 4**), with 23 of 24 samples showing identical calls and only minimal discordance (**Supplementary Figure 2A-B**). The single discrepancy occurred in sample GM19176, where HLA*LA reported C*18:01 and DRAGEN reported C*18:02.

Next, we benchmarked WGS-based HLA*LA typing (WGS+HLA*LA) against clinically validated HLA genotypes generated using GenDX NGSengine (**Figure 1B**). GenDX uses orthogonal PCR-based amplification followed by Illumina sequencing and was therefore treated as the reference truth set. Accuracy was calculated as the proportion of correctly typed alleles per gene relative to the GenDX call. Although HLA*LA and GenDX can profile up to 13 and 10 HLA genes respectively, we restricted the comparison to eight shared loci: Class I (HLA-A, -B, -C) and Class II (HLA-DPA1, -DPB1, -DRB1, -DQB1, -DQA1). Minor Class I loci (HLA-E, -F, -G) were not typed by GenDX at the time of analysis, and DRB3/4/5 reporting was selective; moreover, HLA-DRB4 and HLA-DRB5 are DRB1-linked paralogues with haplotype-dependent presence and are not expected in all individuals (24).

Across all four sequencing platforms (Illumina, MGI, ONT R9, and ONT R10), WGS+HLA*LA achieved consistently high accuracy relative to GenDX, with an overall mean accuracy of approximately 94% (range: 93.7-94.5%; **Figure 2A; Table 2**). Most samples exceeded 95% accuracy, although a small subset, predominantly from ONT datasets, showed reduced performance at select Class II loci (**Figure 2C**). Accuracy was higher for Class I genes (HLA-A, -B, and -C; all >98%) compared with Class II genes, where performance was lower overall and particularly reduced for HLA-DQA1 and HLA-DQB1 (mean 83-85%; **Figure 2B**).

**Figure 2.**
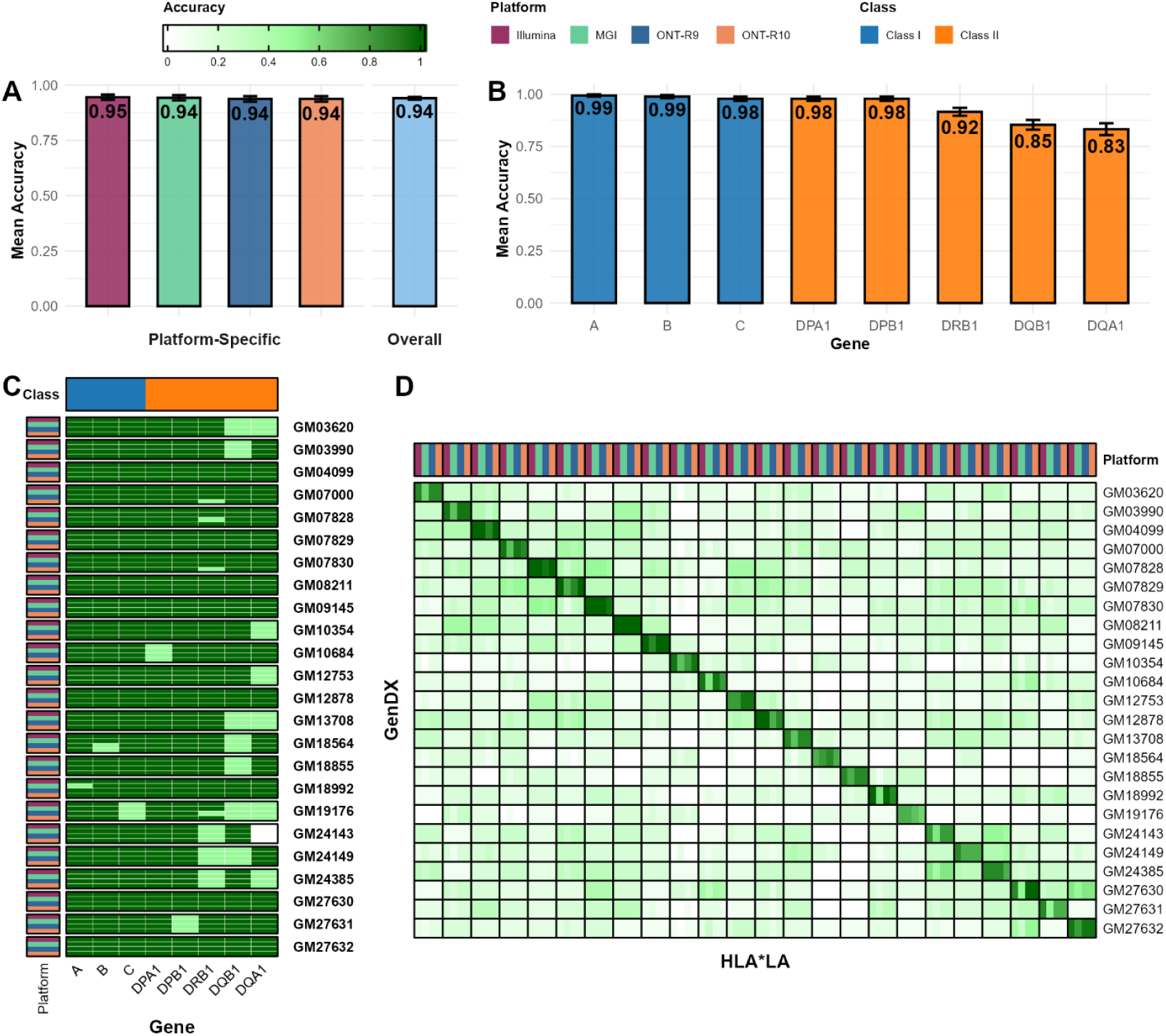
Benchmarking WGS-based HLA genotyping against GenDX across sequencing platforms. **(A)** Mean HLA typing accuracy of WGS+HLA*LA relative to GenDX across sequencing platforms (Illumina, MGI, ONT R9, ONT R10), shown with an overall aggregate. Accuracy was computed as the proportion of correctly typed alleles per locus compared with GenDX reference calls across the shared loci. **(B)** Gene-level accuracy stratified by HLA class, summarizing mean accuracy for Class I (HLA-A, HLA-B, HLA-C) and Class II loci (HLA-DPA1, HLA-DPB1, HLA-DRB1, HLA-DQB1, HLA-DQA1) across platforms. Class I loci showed consistently high accuracy, while reduced performance was primarily observed in specific Class II genes. **(C)** Per-sample, per-gene accuracy heatmap comparing WGS+HLA*LA calls against GenDX for the 24 Coriell cell lines. Each cell represents the allele-level match score for a given locus (1.0 = both alleles match; 0.5 = one allele matches; 0 = no alleles match), using an order-invariant comparison at 2-field resolution. Platform annotations indicate the WGS dataset source for each sample. **(D)** Pairwise concordance matrix comparing HLA*LA-derived genotypes across Coriell samples, illustrating that concordance is highest for matched comparisons and substantially lower for randomly paired mismatched individuals. This permutation-based baseline confirms that the observed agreement is not attributable to chance or shared common alleles.

Inspection of per-sample, per-gene accuracy patterns indicated that departures from perfect accuracy in Class I genes and the DP loci (HLA-DPA1 and HLA-DPB1) were largely driven by isolated discordant specimens (e.g., GM18992 at HLA-A; **Figure 2C**). In contrast, mismatches in other Class II loci were more pervasive across samples, consistent with the known complexity of Class II genotyping and reduced mappability (11) in these regions (**Figure 2C**). These results suggest that while WGS+HLA*LA provides highly reliable typing overall, a subset of Class II loci can remain more challenging to resolve accurately using WGS data alone.

The mean accuracy was highly consistent across sequencing platforms (**Figure 2A**), and mismatches between WGS+HLA*LA and GenDX were observed across all technologies rather than being driven by a single platform (**Figure 2C**). Formal statistical testing supported this observation, with no significant differences detected in platform-level accuracy distributions (paired Wilcoxon signed-rank test: p = 0.233) or mean accuracy values (one-way ANOVA: p = 1.00) (**Methods**). Finally, accuracy values computed from random combinations of mismatched cell lines across platforms were low (permutation median = 0.188, range: 0.125-0.312) and were substantially lower than the observed accuracy (observed median = 0.938, range: 0.500-1.000; one-sided t-test: p = 5.32×10^−20^; **Figure 2D, Supplementary Table 2**), confirming that the high agreement between WGS+HLA*LA and GenDX was not attributable to chance or limited diversity.

Collectively, these results demonstrate that HLA*LA applied to WGS data delivers consistently high HLA genotyping accuracy across sequencing technologies, supporting its use as a robust and platform-agnostic HLA caller in downstream analyses.

### Applicability to biobank samples

The preceding analyses were derived primarily from cell lines, where abundant high-quality DNA can be obtained for WGS. To be useful in real-world biobank settings, WGS-based HLA inference must remain reliable across commonly collected biological samples, including standard blood-derived DNA as well as less invasive sources such as buccal swabs and saliva. We therefore evaluated HLA*LA performance across three sample sources (blood, buccal swab, and saliva) and three sequencing platforms (Illumina, MGI, and ONT R9) (**Figure 1C; Figure 3**).

**Figure 3.**
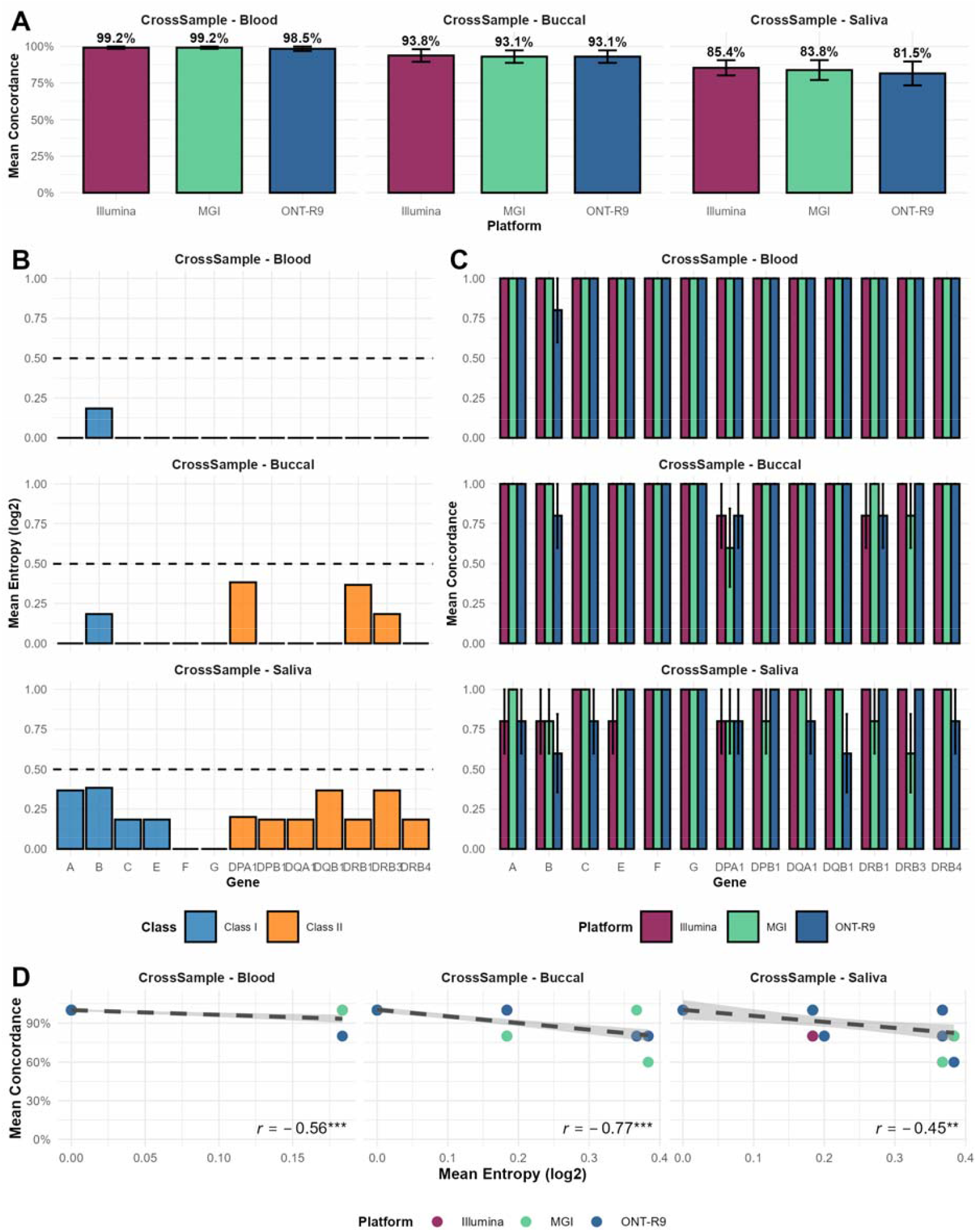
Concordance and entropy of HLA typing across sequencing platforms and sample sources. **(A)** Mean HLA concordance across sequencing platforms (Illumina, MGI, ONT R9) stratified by sample source (blood, buccal swab, saliva). Concordance was computed across all profiled loci using a gene-wise, order-invariant allele matching scheme at 2-field resolution. Error bars represent standard error. **(B)** Mean per-allele entropy (log_2_) across HLA genes for each sample source, stratified by HLA class (Class I: HLA-A, -B, -C, -E, -F, -G; Class II: HLA-DPA1, - DPB1, -DQA1, -DQB1, -DRB1, -DRB3, -DRB4). Higher entropy indicates greater uncertainty in posterior allele assignment. **(C)** Per-gene concordance across platforms for each sample source, highlighting locus-specific sensitivity to sample type and sequencing technology. **(D)** Relationship between mean concordance and mean entropy across platforms within each sample source. Pearson correlation coefficients are shown; shaded regions indicate 95% confidence intervals.

Across sample sources and sequencing technologies, datasets achieved broadly comparable genome-wide coverage (**Table 1**), and HLA*LA produced high-confidence genotypes across most loci. As in our earlier analyses, we harmonized allele comparisons to 2-field resolution for consistency and clinical interpretability. Because orthogonal truth HLA calls were not available for these individuals, we used within-individual concordance across sample sources and platforms as a proxy measure of typing robustness.

Overall concordance was highest for blood-derived DNA across all platforms (98.5-99.2%), followed by buccal swabs (93.1-93.8%) and saliva (81.5-85.4%) (**Figure 3A** and **Supplementary Figure 3A**). Differences between sequencing platforms were most apparent for saliva-derived DNA, where Illumina outperformed MGI and ONT by approximately ∼2% and ∼4%, respectively, whereas cross-platform differences were minimal (<1%) for blood and buccal swab samples (**Figure 3A**). These results indicate that sample source is a stronger determinant of concordance than sequencing platform, with saliva presenting the greatest challenge for consistent HLA inference.

To further evaluate genotype quality, we quantified per-allele entropy (log_2_), a measure of posterior uncertainty in allele assignments returned by HLA*LA. Lower entropy corresponds to higher-confidence calls, while higher values indicate ambiguity. In line with concordance patterns, entropy was lowest for blood-derived DNA (mean = 0.014; median = 0; range = 0-0.918), higher in buccal swabs (mean = 0.086; median = 0; range = 0-1.00), and highest in saliva (mean = 0.214; median = 0; range = 0-1.00) (**Figure 3B**). Concordance and entropy showed a consistent inverse relationship across sample sources (Pearson’s r ranging from -0.45 to -0.77, depending on source; **Figure 3D**), suggesting that uncertainty in posterior allele assignment explains a substantial fraction of reduced concordance in lower-quality sample types. This trend was consistent with observations in the Coriell and SGDP analyses (**Supplementary Figure 4**). Across datasets, entropy was generally lower for Class I loci than for Class II loci, with the largest uncertainty often observed in *HLA-DQA1* and *HLA-DQB1* (**Figure 3B-C**).

Together, these results support the applicability of WGS+HLA*LA to real-world biobank specimens, while highlighting that buccal and saliva-derived DNA—particularly saliva—can introduce increased uncertainty and reduced cross-platform concordance, most prominently in Class II loci.

### Robustness to reduced sequencing coverage for increased cost-effectiveness

The analyses performed up to this point used ∼30x WGS datasets, a typical depth for clinical and research sequencing (25). Cost-effective HLA typing applications will benefit from reduced sequencing, which requires less DNA, lowering sequencing costs and shortening turnaround time without hopefully compromising genotyping accuracy. To quantify the impact of reduced coverage, we downsampled five Coriell cell line datasets across platforms to 25x, 20x, 15x, 10x, and 5x and compared inferred HLA genotypes to each sample’s 30x HLA*LA calls as the within-sample reference.

We confirmed that genome-wide down-sampling proportionally reduced coverage in the HLA region (**Figure 4A**). WGS-based HLA genotypes remained highly consistent down to 15x across platforms (**Figure 4B**, 2-field). At 15×, two-thirds of sample–platform combinations (13/20) achieved perfect concordance with their corresponding 30× calls, and all remained above 90% concordance (**Figure 4C**). Below 15x, accuracy declined more markedly but remained >85% after a two-thirds reduction in coverage. Concordance at 1-field resolution stayed >90% even at 5x, whereas 3-field predictions were substantially more sensitive to reduced coverage, reflecting the difficulty of resolving long, polymorphic HLA sequencies (**Figure 4B**). Overall, Illumina, followed by ONT-R10 consistently outperformed the ONT R9 and MGI (**Figure 4B**).

These results demonstrate that WGS⍰based HLA typing remains highly accurate down to 15x coverage, supporting substantial reductions in sequencing depth without sacrificing performance significantly. This establishes a practical lower⍰coverage threshold that can markedly improve cost⍰effectiveness for large⍰scale biobank and population genomics studies.

**Figure 1.**
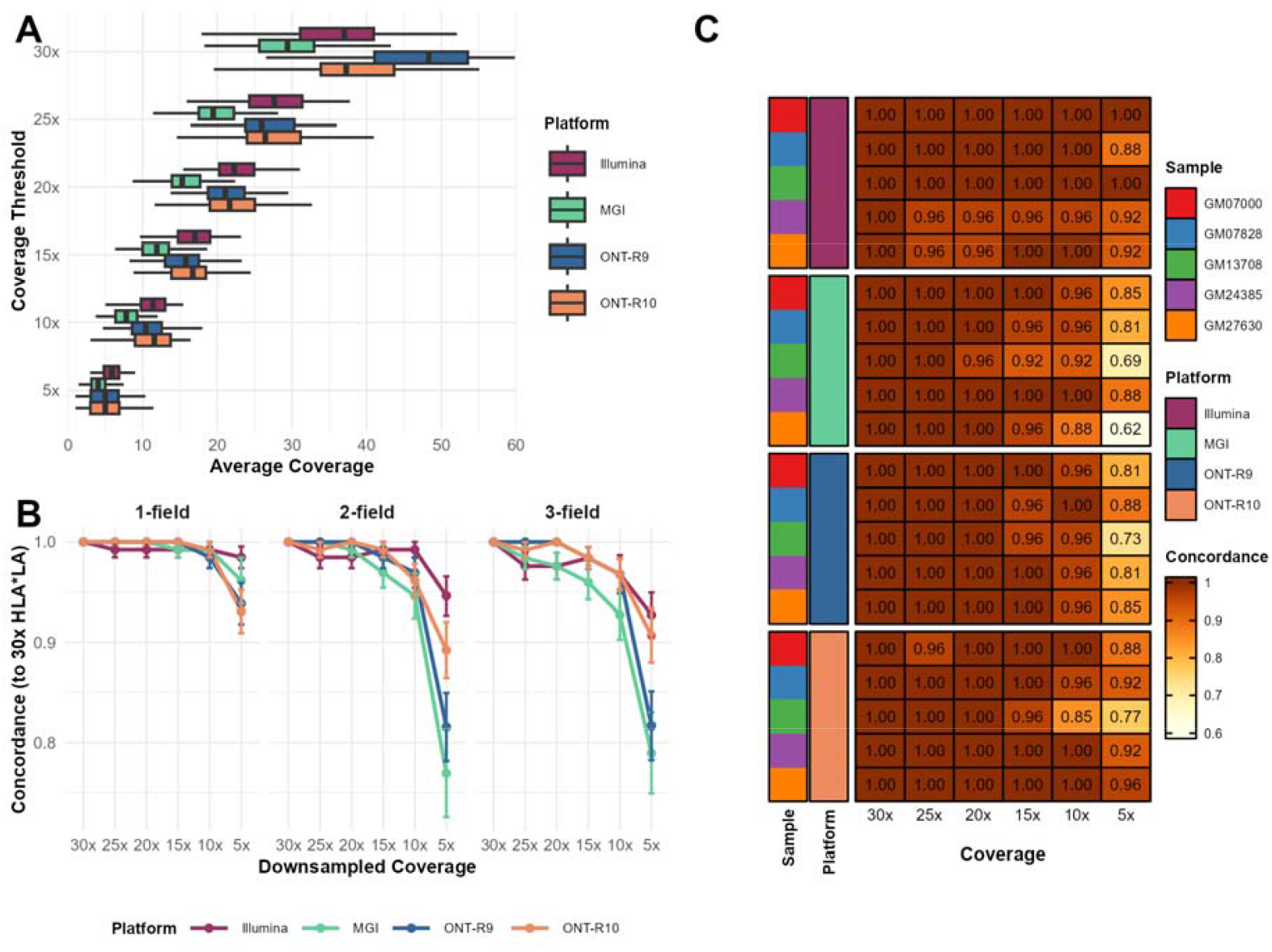
WGS-based HLA genotypes robustness to reduced sequencing depth. **(A)** Distribution of average per-sample HLA gene coverage across sequencing platforms and downsampling thresholds. Boxplots represent the observed average coverage for each platform at the intended threshold (5x to 30x), confirming successful and consistent downsampling across samples. **(B)** Concordance of HLA typing results relative to the 30x HLA*LA calls at 1-field, 2-field, and 3-field resolution. Accuracy declines with reduced coverage, particularly below 15x and most markedly at 3-field resolution. Illumina, MGI, and ONT platforms show similar performance trends. **(C)** Heatmap of sample-level HLA typing concordance to 30× across downsampled coverage levels and platforms. Each cell reflects the mean accuracy across all loci for a given sample-platform-coverage combination. High concordance (≥0.96) is observed in most cases down to 10x, with some degradation at 5x, particularly for ONT-R9.

## Discussion

In this work we evaluated the applicability of WGS data to accurately genotype 13 major HLA genes (**Figure 1**). By integrating WGS datasets generated from multiple biospecimen types, we systematically assessed the impact of DNA sources, sequencing technology, and coverage on HLA typing performance. In addition to evaluating concordance across platforms, we benchmarked WGS-based calls against a clinical-grade reference assay (GenDX). Overall, WGS-based HLA genotyping achieved ∼95% accuracy across sequencing technologies (**Figure 2A; Table 1**), with performance remaining above 95% even at half coverage (**Figure 4B**). Concordance was similarly high for cell lines and real-world blood and buccal swab samples, whereas saliva-derived samples showed reduced performance (**Figure 3**). Together, these findings support the feasibility of accurate HLA typing directly from WGS data across common biospecimen types.

An important question is why one would use genome-wide sequencing to type the relatively compact ∼3-5 Mb HLA region (**Figure 1**). While targeted assays remain cost-effective for single-purpose HLA typing—such as PCR-based or amplicon-driven platforms (e.g. GenDX), (26–29), they rely on locus-specific amplification and therefore fix the set of profiled genes and remain prone to primer bias and allele dropout (30–34). As large-scale biobanks (35–38) increasingly generate whole-exome or -genome sequencing data for diverse applications, the ability to infer HLA from existing datasets becomes advantageous. For example, UK Biobank and Estonian Biobank data have already been used to study HLA diversity and disease associations (36,39). Therefore, the capability to accurately type HLA from WGS greatly increases dataset reusability and facilitates population-scale HLA registry development, which may improve donor-recipient matching in transplantation.

Our study also provides a benchmark comparing long-read Nanopore sequencing with the market-leading Illumina platform. Despite higher error rates, ONT achieved HLA genotyping performance similar to Illumina (**Figure 2A**), with Illumina showing only marginal advantages for blood and buccal swabs and more noticeable differences in saliva and at <15× coverage (**Figure 3** and **Figure 4**). Although not tested in this work, adaptive sampling could be also useful for HLA typing if WGS data is not of interest. Using e.g. a minION together with buccal swaps could indeed be a cost-efficient broad method independent of sequencing centers. The SRS-based MGI platform performed similarly to Illumina and ONT. Taken together, these results demonstrate consistent cross-platform performance, supporting the integration of multi-technology WGS datasets for HLA studies that is an increasingly common need as biobanks diversify their sequencing pipelines.

Even when HLA typing is the primary goal, WGS offers advantages over targeted approaches. Typically, targeted assays sequence HLA genes (or individual exons) separately, constrain which loci can be analyzed and limiting interpretation as interest grows in historically overlooked or newly relevant genes. For instance, many earlier tools disproportionately optimized Class I loci (*HLA*⍰*A*, ⍰*B*, ⍰*C*) due to their simpler structure and stronger historical clinical focus (40), whereas WGS enables analysis of all classical and non⍰classical genes as software capabilities expand (e.g., HLA*LA, updated DRAGEN versions). Moreover, LRS technologies like ONT and PacBio provide not only longer fragments that increase HLA typing accuracy and resolution but also enable long-range phasing and methylation analysis (41). Thus, WGS provides a future⍰proof substrate for broader HLA interrogation.

We also evaluated minimally invasive sampling strategies for scalable clinical or consumer applications. Buccal swabs achieved accuracy comparable to blood and cell lines (**Figure 2** and **Figure 3**), despite yielding less and more fragmented DNA, demonstrating that WGS-based HLA typing is robust to lower DNA quality. By contrast, saliva showed ∼15% reduced accuracy (**Figure 3**), highlighting limitations for certain self-collection methods. Additionally, we examined how much sequencing is required to maintain high accuracy: 15× coverage achieved >95% accuracy at 2-digit resolution, 10× sufficed for 1-digit typing and >=20× was needed for 3-digit resolution (**Figure 4**).

Our study has limitations. Most datasets were derived from Coriell reference cell lines, and our real⍰world evaluation included only five individuals, although profiling multiple DNA sources and platforms provided complementary diversity. While our samples span varied ancestries, they do not capture the full extent of HLA variation, and accuracy in other studies may differ as more alleles and populations are included. There are other HLA callers other than HLA*LA which we have not evaluated; our primary goal though was to demonstrate the accuracy of WGS-based HLA typing more than benchmarking different tools. Finally, Class II genotyping, particularly for DQB1 and DQA1, remains challenging (**Figure 2** and **Figure 3**), underscoring the need for improved calling performance and additional high-quality references.

## Conclusions

This study demonstrates that WGS can deliver accurate, platform-agnostic HLA genotyping across diverse sample types and sequencing depths. Through validation against a clinical reference assay and systematic evaluation of coverage and resolution trade-offs, we establish practical thresholds for reliable HLA inference without targeted enrichment. These results support the use of WGS as a scalable substrate for population-scale immunogenomics, registry development, and future clinical integration, while maximizing the value of existing and emerging biobank datasets.

## Methods

### Simons Genome Diversity Project

Five individuals were selected at random from the Simons Genome Diversity Project (SGDP), a high-coverage whole-genome sequencing resource spanning globally diverse ancestry. Publicly available processed data files aligned to GRCh38 were obtained from the associated data repository (42). HLA typing was performed in-house using HLA*LA (v1.0.4) with default parameters and the provided population reference graph.

### Coriell reference samples

#### Cell line culture

Twenty-four lymphoblastoid cell lines (Supplementary Table 7) were purchased from the Coriell Cell Repositories (Camden, NJ, USA) and cultured according to the supplier’s instructions. We selected live cell cultures rather than DNA reference materials to preserve the native DNA structure, as transportation and storage of purified DNA may lead to fragmentation that could compromise the long reads required for Oxford Nanopore Technologies (ONT) sequencing. Upon receipt, cells were supplied in T-12.5 ml tissue culture flasks and incubated overnight at 37 °C. The following day, cultures were transferred to 50 ml centrifuge tubes and centrifuged at 100 × g for 10 min. The supernatant was discarded, and the pellet was resuspended in complete growth medium (RPMI 1640 supplemented with 15% fetal bovine serum). Cells (∼1 × 10^6^) were seeded into 25 ml tissue culture flasks containing 10 ml of medium and incubated at 37 °C with 5% CO_2_. Cultures were harvested upon reaching adequate density, and viable cell counts were determined.

#### DNA extraction

Genomic DNA was extracted using the PureLink™ Genomic DNA Mini Kit (Cat. No. K182001, Thermo Fisher Scientific) according to the manufacturer’s protocol. Briefly, cell pellets were obtained by centrifugation at 15,000 rpm for 5 min, followed by removal of the growth medium. Pellets were resuspended in 200 µl phosphate-buffered saline (PBS), and 20 µl each of Proteinase K and RNase A were added. After thorough mixing, 200 µl of PureLink Genomic Lysis/Binding Buffer was added, and the lysate was vortexed to homogeneity and incubated at 55 °C for 10 min.

An equal volume (200 µl) of 96–100% ethanol was then added, and ∼640 µl of the lysate was transferred to a spin column and centrifuged for 1 min. The flow-through was discarded, and the column was washed sequentially with 500 µl of Wash Buffer 1 and 500 µl of Wash Buffer 2. DNA was eluted with 200 µl of elution buffer by centrifugation at maximum speed for 1 min at room temperature.

### Sequencing

#### Illumina

Whole-genome sequencing (30x) libraries were prepared using the Illumina® PCR-Free Prep kit, following the manufacturer’s instructions. Genomic DNA input (250–750 ng) was fragmented using bead-linked transposomes, and ligation was performed with IDT® for Illumina® UMI DNA/RNA UD Indexes Set A (96 indexes, 96 samples). Libraries from 24 samples were pooled based on index compatibility and sequenced on the NovaSeq 6000 platform using the NovaSeq 6000 S4 Reagent Kit v1.5 (300 cycles).

#### MGI

Libraries were prepared with the MGIEasy PCR-Free DNA Library Prep Kit (96 RXN; Cat. No. 1000013457, MGI, Shenzhen, China) using 900 ng of DNA in 48 µl. Preparation steps included fragmentation, size selection, end repair, adapter ligation, denaturation, circularization, and exonuclease digestion. For double-size selection, DNA EasyClean beads were applied at 0.6× and 0.2× ratios. Library quality was assessed using the Qubit™ ssDNA Assay Kit, with concentrations of 0.6–3 ng/µl considered suitable for DNA nanoball (DNB) preparation. DNB concentrations of 8–40 ng/µl were pooled and loaded onto the DNBSEQ-T10RS platform, using the DNBSEQ-T10RS DNB Sequencing Set (FCL PE100; Cat. No. 940-000078-00). Sequencing was performed on the DNBSEQ-T10 flow cell, and raw data were processed with ZLIMS Elite v1.0.5.2 using the MEGABOLT_2 pipeline.

#### ONT

Libraries were prepared using the ONT Ligation Sequencing Kit 114 (Cat. No. SQK-LSK114-XL, ONT, Oxford, UK) with 1000 ng of DNA in 50 µl. Preparation steps included normalization, mechanical fragmentation (FastPrep), end repair, and adapter ligation. Library quality was assessed using the Qubit™ dsDNA Assay Kit, with ≥400 ng/µl used for loading onto the PromethION flow cell. Sequencing was performed on the PromethION 48 system. The recorded data was analyzed using MinKNOW software with Dorado.

### Primary and secondary analysis

#### Illumina

Raw BCL files were processed using the on-premises Illumina DRAGEN Germline Pipeline v4.1.7 to perform demultiplexing, basecalling, and FASTQ generation. DRAGEN preprocessing included adapter trimming and quality filtering, followed by alignment to the GRCh38 reference genome. Variant calling (SNPs, indels, CNVs, STRs, HLA) was performed using DRAGEN’s integrated callers. Outputs included CRAM (alignment) and VCF/gVCF (variants) files, with summary statistics and log files archived for each sample.

#### MGI

Native CAL files were processed on the on-premises MGI Ztron Pro platform for demultiplexing and basecalling to FASTQ format. Reads underwent adapter trimming and quality filtering before alignment to GRCh38. Variant calling (SNPs, indels) was performed with ZTRON Pro. CNV detection was carried out using CANVAS v1.40.0.1613, STR calling with ExpansionHunter v5.0.0, and HLA typing with HLA-LA v1.0.3. Outputs (CRAM and VCF/gVCF) were further processed in a custom in-house pipeline on the G42 Cloud. Summary statistics and processing logs were archived.

#### ONT

Native FAST5 files from the PromethION P48 system were processed on a GPU-enabled environment (NVIDIA A100) to perform demultiplexing and basecalling to FASTQ/uBAM format. An in-house ONT analysis pipeline on G42 Cloud merged per-run uBAM files using Samtools v1.19, performed QC with Fastp v0.23.4, and aligned reads to GRCh38 using Sentieon-accelerated Minimap2 v2.22. Alignment QC was conducted using Alfred v0.2.6.

Variants were called with Clair3 v1.0.4 (SNVs/indels) and Sniffles2 v2.2 (SVs). CNVs were detected with Spectre v0.1.1.alpha, STRs with Straglr v0.2.4, and HLA typing with HLA-LA v1.0.3. SMN1/2 copy number analysis was performed with Hapdup v0.12.3 / Hapdiff v0.8.8. Variant QC was conducted with VariantQC, and outputs (CRAM, VCF/gVCF) along with software logs and summary statistics were archived.

### Down-sampled datasets

To evaluate the impact of sequencing depth on HLA typing accuracy, we selected five Coriell lymphoblastoid cell line samples (GM07000, GM07828, GM13708, GM24385, GM27630) for silico down-sampling. These samples were chosen to represent high-quality, full-coverage datasets across platforms, ensuring that any differences observed after down-sampling would be attributable to reduced coverage rather than pre-existing sequencing or alignment artefacts. Platform-specific aligned read files (BAM or CRAM format) were down-sampled using SAMtools (v1.20) with the -s parameter to randomly subsample reads while preserving read pairing where applicable. Target average mapping coverages were set at 5x intervals from 5x to 30x (i.e., 5x, 10x, 15x, 20x, 25x, and 30x). The original full-coverage datasets served as the 30x baseline for comparison. Down-sampled files were subsequently processed using the same HLA typing workflow as full-coverage datasets, ensuring that all other parameters remained constant. This approach enabled direct quantification of accuracy loss, locus, and resolution-dependent performance across decreasing sequencing depths.

### Blood, buccal swab and saliva samples

Blood, buccal, and saliva samples were obtained from five healthy volunteers at the time of the study, all of whom were employees of M42. Recruitment followed M42’s established recruitment and consent policies, ensuring a fair and transparent approach to participant selection. Convenience sampling was used, drawing from a pool of accessible volunteers who were willing to participate, thereby maintaining both practicality and fairness in the sampling method. Each participant provided all three sample types (saliva, buccal, and blood). Written informed consent was obtained from all participants, outlining their rights, as well as the intended use and storage of the samples. The collected samples were subsequently processed and sequenced using the same laboratory and bioinformatics workflows as applied to the Coriell cell line data.

### HLA Typing

In all datasets, HLA genotyping was performed using HLA*LA (v1.0.3) (8), a graph-based HLA caller designed to resolve highly polymorphic HLA loci from whole-genome sequencing data. HLA*LA operates by aligning sequencing reads to a population reference graph constructed from the IMGT/HLA database, followed by assembly-based genotyping to predict phased alleles. By default, HLA*LA produces genotype calls for the classical Class I genes (HLA-A, HLA-B, HLA-C), non-classical Class I genes (HLA-E, HLA-F, HLA-G), and Class II genes (HLA-DQA1, HLA-DQB1, HLA-DRB1, HLA-DPA1, HLA-DPB1), returning two alleles per locus to represent diploid genotypes. Each genotype call is accompanied by a posterior probability score that reflects the confidence of the allele assignment; low-confidence calls (e.g., <0.5 posterior probability) were flagged for further inspection. For the Coriell cell line datasets sequenced on Illumina, we additionally ran the Illumina DRAGEN HLA caller (v4.1.7) (7), which outputs genotypes for HLA-A, -B, and -C only. DRAGEN uses a k-mer–based alignment and likelihood-scoring approach optimized for speed, producing two-field resolution genotypes with associated quality metrics. DRAGEN calls for these three loci were compared directly to HLA*LA calls for cross-validation.

### HLA Typing Accuracy and Concordance Analysis

#### Ground Truth Set and Accuracy Calculation

To evaluate HLA typing accuracy, we used a reference set of high-confidence HLA calls generated using the GenDX NGSengine pipeline on the Coriell panel of genomic DNA samples. These calls served as ground truth for benchmarking across sequencing platforms and tissues.

Accuracy was assessed at two-field resolution (e.g., HLA-A*01:01) for HLA genes: HLA-A, -B, -C, DQA1, DQB1, DRB1, DPA1, and DPB1. For each gene, allele calls from the test platform were compared against the corresponding GenDX reference alleles.

Alleles were treated as unordered pairs per gene. A match was considered valid if the called allele set matched the ground truth regardless of allele order (e.g., {01:01, 02:01} matched {02:01, 01:01}). Gene-level accuracy scores were assigned as follows: 1.0 if both alleles matched the GenDX reference, 0.5 if one allele matched, and 0.0 if neither allele matched. Gene calls that were missing or ambiguous were excluded from downstream accuracy calculations.

Per-sample accuracy was then computed as the mean gene-level accuracy score across all evaluable genes, yielding a continuous accuracy metric ranging from 0 to 1:

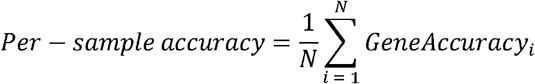

where N denotes the number of genes with valid calls for that sample. Accuracy summaries were generated by platform, gene, and sample type and served as the basis for downstream statistical comparisons.

#### Accuracy Distribution and Mean Differences Testing

To assess whether observed differences in accuracy reflected genuine sequencing platform effects rather than stochastic variation, we performed two complementary statistical analyses of per-sample gene-level accuracy scores. All analyses were conducted independently for each HLA gene and accounted for the paired study design, in which the same Coriell samples were sequenced across multiple platforms.

First, we applied pairwise Wilcoxon signed-rank tests to compare the distributions of per-sample gene-level accuracy scores between platforms (Supplementary Table 1). This non-parametric approach is well-suited for bounded accuracy values (0, 0.5, 1.0) and explicitly accounts for the paired nature of the data by comparing matched samples across platforms. Comparisons labeled as “Not tested” reflect cases where accuracy scores were identical across platforms or lacked sufficient variability to permit statistical evaluation. P-values were adjusted for multiple testing using the Benjamini–Hochberg procedure.

Second, we performed a one-way analysis of variance (ANOVA) followed by Tukey’s Honest Significant Difference (HSD) post-hoc testing to evaluate whether mean gene-level accuracy differed across platforms. This parametric analysis provides a complementary perspective to the Wilcoxon tests by directly testing for differences in average performance while accounting for within-gene variance across platforms. Confidence intervals and adjusted p-values were used to assess statistical significance.

Across all genes and platforms, neither approach identified statistically significant platform-specific differences in accuracy after multiple testing correction, indicating that WGS-based HLA typing accuracy was highly consistent across sequencing technologies.

#### HLA Concordance Analysis

To assess intra- and inter-platform reproducibility of HLA typing independently of an external reference, we performed pairwise concordance analyses across samples typed on multiple sequencing platforms or derived from different tissue sources. In contrast to accuracy analyses, which benchmarked calls against a clinical ground truth, concordance evaluates the agreement between HLA calls obtained from different datasets for the same individual.

For each pairwise comparison, concordance was computed at the gene level by assessing whether the same alleles were called between two samples for a given HLA locus. Alleles were treated as unordered pairs, and concordance was defined as:

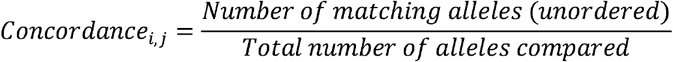

resulting in concordance scores of 1.0 (both alleles matched), 0.5 (one allele matched), or 0.0 (no alleles matched) for diploid loci.

Pairwise concordance matrices were generated using all samples with shared identifiers (e.g., the same individual or Coriell cell line) (Supplementary Tables 2-6). For cross-platform comparisons, samples with HLA calls from at least two platforms were included (e.g., Illumina vs MGI, Illumina vs ONT-R9, ONT-R9 vs ONT-R10). For cross-tissue comparisons, matched blood, buccal, and saliva samples were compared within each individual.

Samples with missing calls or unresolved genotypes at a given locus were excluded from concordance calculations for that gene. Gene-level concordance scores were then aggregated across loci to compute an overall per-sample agreement rate. Concordance results were visualized using heatmaps generated with the ComplexHeatmap R package, with matrices faceted by sequencing platform or tissue source to highlight reproducibility patterns across technologies and biospecimen types.

### Entropy-Based Analysis of HLA Typing Discordance

To assess whether allele-level uncertainty was associated with discordance in HLA typing across sequencing platforms, we performed an entropy-guided concordance analysis using high-resolution HLA typing results. For each sample, gene, and platform combination, allele-level uncertainty was summarized using Shannon entropy (log_2_ scale), computed from posterior probabilities of HLA allele assignments (see HLA typing and entropy quantification).

#### Normalization of Allele Sets and Definition of Discordance

To enable consistent pairwise comparisons independent of allele order, each biallelic HLA call was normalized into a sorted, unordered allele set (e.g., “*01:01;*02:01” equivalent to “*02:01;*01:01”). Inter-platform discordance was defined as instances where a sample had differing normalized allele sets for the same gene across sequencing platforms. Intra-platform discordance was assessed analogously for samples with repeated calls from the same platform and gene (e.g., technical replicates).

#### Association Between Entropy and Discordance

To assess whether allele-level uncertainty was associated with discordance in HLA typing across sequencing platforms, we examined the relationship between per-sample entropy and inter-platform concordance. For each sample–gene pair, HLA allele calls were normalized into unordered allele sets and classified as concordant or discordant based on agreement across platforms.

Per-sample mean entropy values were then compared with discordance status to evaluate whether higher uncertainty was associated with reduced reproducibility. Associations were quantified using Spearman rank correlation, a non-parametric measure robust to bounded accuracy values and non-normal distributions.

Across datasets with sufficient variability, higher entropy was consistently associated with increased discordance between platforms (**Figure 3D**), indicating that allele-level uncertainty captured by entropy reflects reduced stability of HLA calls. In datasets where discordance events were rare or entropy values exhibited limited variance (e.g., CrossSample–Blood), correlation estimates were unstable and therefore not reported.

These results support entropy as a meaningful indicator of HLA typing reliability across sequencing technologies, rather than a platform-specific artifact.

### Downsampling-based concordance analysis across platforms

#### Read downsampling and achieved coverage

For each sample–platform, reads were **randomly subsampled** to target genomic depths of **30×, 25x, 20x, 15x, 10x, and 5x** using probabilistic read selection with a fixed seed to ensure reproducibility. The sampling fraction was chosen as (*target/observed coverage*). After subsampling, BAMs were reindexed and **achieved coverage** was verified by computing average depth over the HLA region; the distributions shown in **Figure 4A** confirm that achieved depths closely matched targets across platforms.

#### HLA typing and resolution standardization

HLA genotypes were called on each downsampled dataset using **HLA*LA** (reference), yielding two alleles per gene when callable and posterior probabilities per allele. Calls were compared at **1, 2, and 3 field** resolution by truncating allele labels to the desired field depth (e.g., HLAA*02:01:01 → 1-field 02, 2-field 02:01, 3-field 02:01:01). Genes with known structural variation (e.g., **DRB3/4/5**) were allowed to be **monoallelic;** in such cases the denominator for accuracy calculations equals the number of expected alleles present (1 instead of 2).

#### Concordance to 30× calls (reference truth)

Per sample and resolution, 30× HLA*LA calls were treated as the within sample reference. For each downsampled depth, per gene accuracy was defined as the fraction of reference alleles matched (0, 0.5, or 1.0 for diploid loci; 0 or 1.0 for monoallelic loci). Per sample concordance equals the mean of per gene accuracies across all loci typed in both conditions. Panel **Figure 4B** reports mean concordance by platform and coverage; **Figure 4C** shows the per sample heatmap.

#### Handling missing and ambiguous calls

Alleles without confident HLA*LA support were recorded as missing and excluded from the numerator for accuracy but retained in the denominator for callrate. For loci known to be absent in some haplotypes (e.g., DRB paralogs), presence/absence was first determined from 30× calls; denominators were adjusted accordingly to avoid penalizing biologically absent alleles. Multiallelic or ambiguous outputs were resolved at the target field by string match after truncation; ties were treated as incorrect unless one of the returned alleles exactly matched the reference at that resolution.

### Plotting

Concordance metrics and heatmaps were generated using R. Heatmaps were generated using the ComplexHeatmap R package. ROC analysis and plots were generated with PRROC and ggplots R packages.

## Supporting information

Supplementary Table 7 - HLA output

## Data Availability

The sequencing data generated in this study have been deposited in the database of Genotypes and Phenotypes (dbGaP) under controlled access (Accession Number: phs004346).

## Abbreviations

HLA: Human leukocyte antigen
HTS: High-throughput sequencing
LRS: Long-read sequencing
Mb: Megabases
MHC: Major histocompatibility complex
ONT: Oxford Nanopore Technologies
PCR: Polymerase chain reaction
SRS: Short-read sequencing

## Declarations

### Ethics approval and consent to participate

This study involved both publicly available human cell lines and newly collected human biospecimens. This study used samples (GM03620, GM03990, GM04099, GM07000, GM07828, GM07829, GM07830, GM08211, GM09145, GM10354, GM10684, GM12753, GM12878, GM13708, GM18564, GM18855, GM18992, GM19176, GM24143, GM24149, GM24385, GM27630, GM27631, GM27632) from the Coriell Institute for Medical Research. The collection and use of blood, buccal swab, and saliva samples from five adult participants were approved by the M42 Research Ethics Committee under protocol number HCRC/24/2024/1. No animal data or animal-derived materials were used in this study.

### Consent for publication

The study and this publication were approved by Medical Research and Development committee from the Department of Health – Abu Dhabi under the protocol number DOH/ADHRTC/2025/599.

### Authors’ contributions

C.C. conceived and designed the study, performed analyses, and wrote the manuscript. J.Q. contributed to study design, analysis, and manuscript revision. A.A.Y.A.S., A.E.-K., A.A., C.M., F.A., G.K., H.W., H.S., J.M., O.S., S.E., S.P., V.G., D.G., H.S., S.B., and T.C. contributed to data generation, quality control, and interpretation. F.S. and L.P. provided input on study design, analytical approach, and interpretation. T.M. supervised the project and contributed to manuscript development. All authors reviewed and approved the final manuscript.

## Acknowledgements

We thank our colleagues at M42 who contributed to this work but are not listed as authors, as well as Jawahar and Rich Carter from Oxford Nanopore Technologies for their technical guidance and support.

## Supplementary Results

### Independent validation of HLA*LA performance in the SDGP dataset

Across the three sequencing platforms, genome-wide coverage was comparable (**Supplementary Figure 1D**), and HLA*LA produced high-confidence genotypes across all 13 HLA loci, including at G-group resolution (**Supplementary Figure 1E**). Because HLA*LA reports alleles at G-group resolution (3-field), we harmonized all downstream analyses to 2-field resolution, which captures amino-acid–level polymorphism and is widely used in clinical interpretation (21–23). To quantify cross-platform concordance, we compared allele calls for the same individual across sequencing technologies using a gene-wise, order-invariant matching scheme. For each locus, concordance was scored 1.0 if both alleles matched, 0.5 if one allele matched, and 0 if neither allele matched. Loci with missing or incomplete calls in either platform were excluded from the denominator for that comparison.

Overall, HLA*LA exhibited high cross-platform reproducibility in SGDP, with a mean per-sample concordance of 93.9% (range: 88-100%; **Supplementary Figure 1A-C**). Concordance between datasets from the same individual was consistently higher than concordance between different individuals (**Supplementary Figure 1A, Supplementary Table 3**), indicating that shared common alleles did not artificially inflate agreement. At the locus level, approximately half of the profiled genes (including HLA-A, HLA-DQB1, HLA-DRB1, HLA-DPA1, HLA-E, and HLA-F) achieved 100% concordance across all platform pairings (**Supplementary Figure 1B**). Concordance for the remaining loci generally exceeded 78%, with the lowest agreement observed for HLA-G in Illumina versus long-read comparisons (< 72%.). Importantly, we did not observe a clear relationship between lower-concordance loci and either gene-level coverage (**Supplementary Figure 1D**) or the HLA*LA genotype confidence score (**Supplementary Figure 1E**) (7).

## Supplementary Figures

**Supplementary Figure 1.**
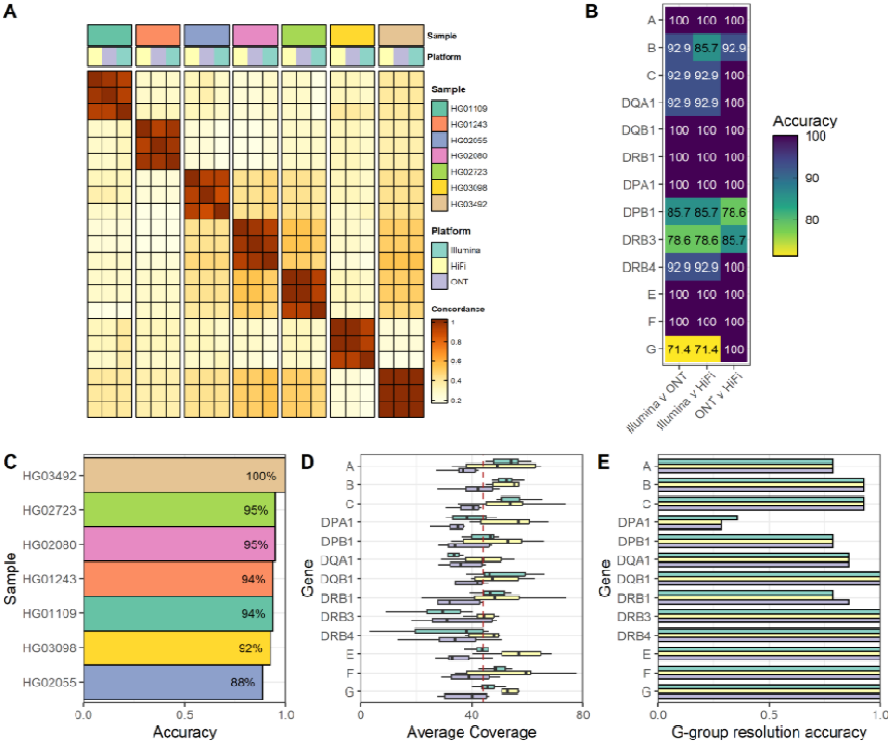
HLA*LA produces HLA calls consistent across sequencing technologies in the SGDP dataset. (A) Pairwise per-sample HLA concordances. (B) Concordance of HLA allele calling across genes per platform comparison. (C) HLA calling accuracy per sample. (D) Per gene allele coverage cross platforms. (E) Percentage of HLA allele calls across samples with HLA*LA’s quality score (Q1) of 1. Q1 approximates the probability of the HLA call for each allele in each sample and ranges from zero to one.

**Supplementary Figure 2.**
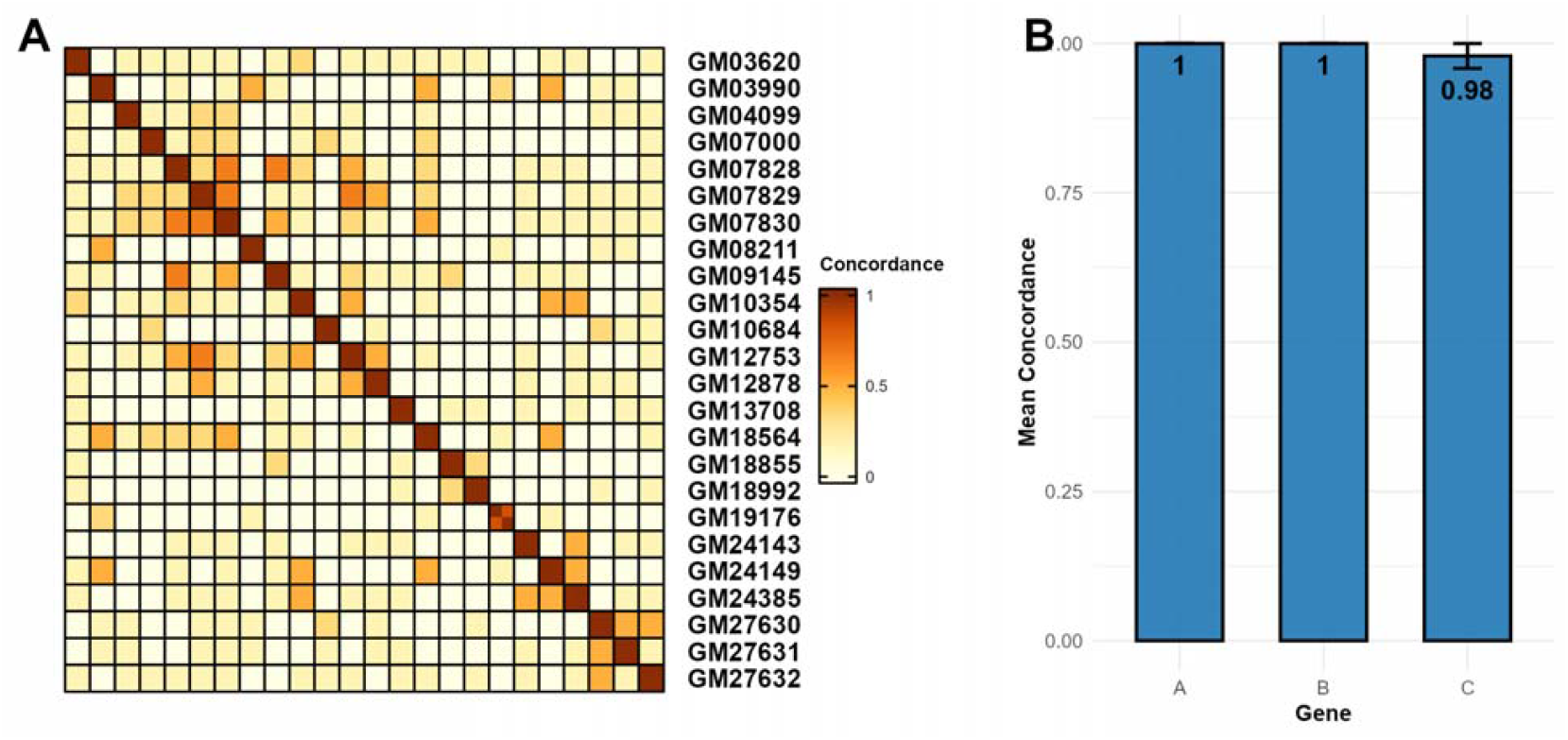
Concordance between DRAGEN and HLA*LA callers. **(A)** Pairwise concordance across 24 Coriell samples sequenced on Illumina. **(B)** Overall per-gene concordance.

**Supplementary Figure 3.**
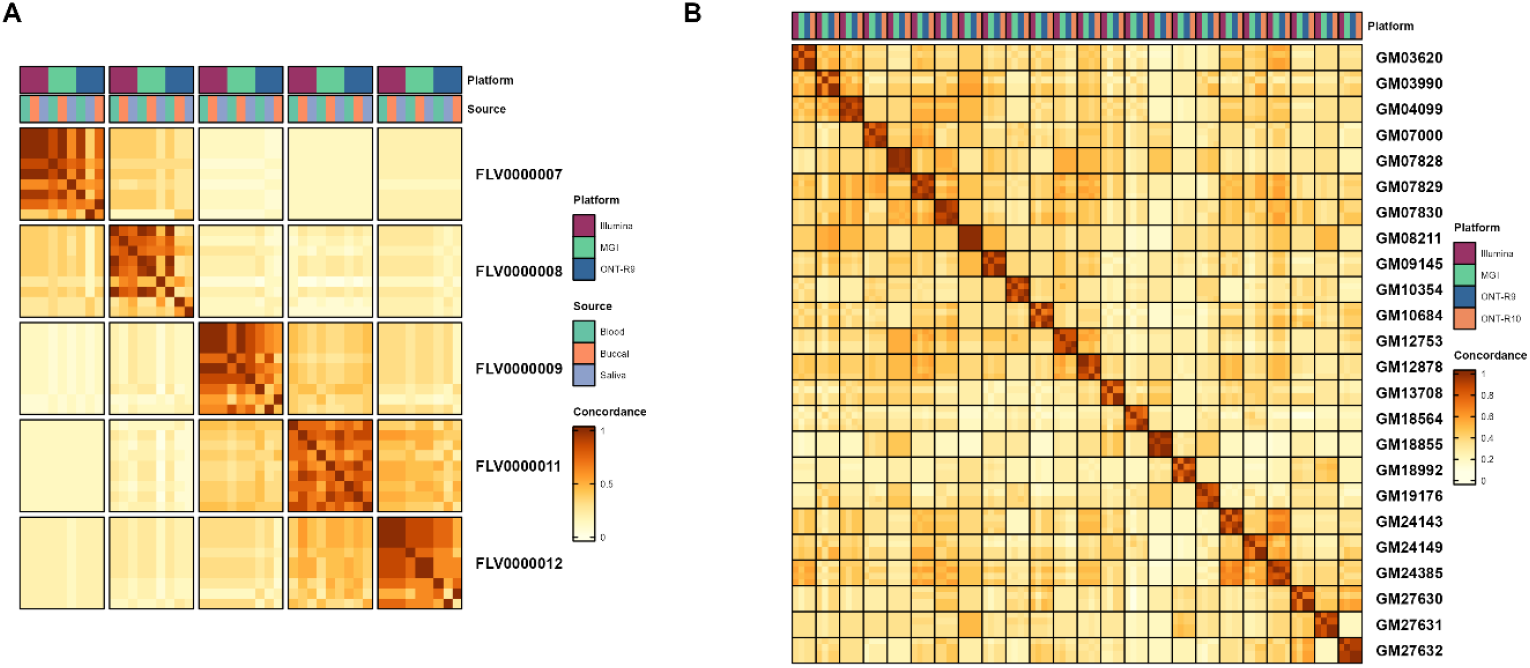
Concordance across Cross-Sample and Coriell datasets. **(A)** Cross-sample concordance across blood, buccal, and saliva samples per individual, sequenced on three platforms. **(B)** Cross-platform HLA concordance heatmap across Illumina, MGI, ONT-R9, and ONT-R10 for Coriell samples.

**Supplementary Figure 4.**
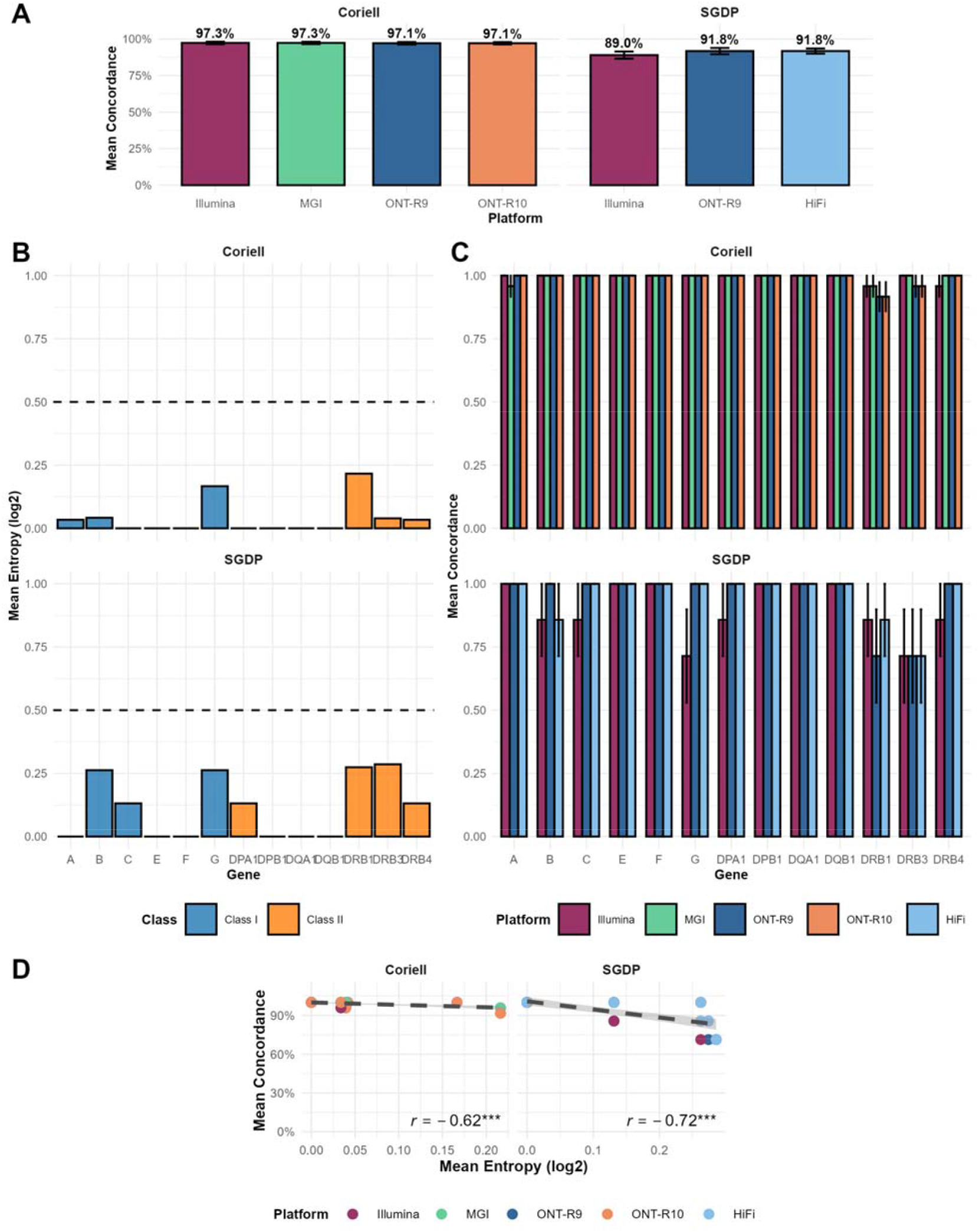
Concordance and entropy of WGS-based HLA typing across datasets. **(A)** Mean HLA concordance across sequencing platforms for the Coriell and SGDP datasets. Concordance was computed across all profiled loci using a gene-wise, order-invariant allele matching scheme at two-field resolution. Bars indicate mean concordance per platform; error bars represent standard error. **(B)** Mean allele-call entropy (log_2_) per HLA gene, stratified by dataset and HLA class (Class I: HLA-A, -B, -C, -E, -F, -G; Class II: HLA-DPA1, -DPB1, -DQA1, -DQB1, -DRB1, -DRB3, -DRB4). Entropy reflects variability in allele assignment across calls, with higher values indicating greater ambiguity. **(C)** Per-gene concordance across sequencing platforms within each dataset. While most loci show near-perfect concordance across technologies, select Class II genes exhibit increased variability, particularly in the SGDP dataset. **(D)** Relationship between mean per-gene concordance and mean entropy across platforms within each dataset. Each point represents a platform–gene combination. A strong inverse correlation is observed in both Coriell (*r* = −0.62) and SGDP (*r* = −0.72), indicating that loci with higher entropy tend to show reduced cross-platform concordance. Shaded regions denote 95% confidence intervals.

## Supplementary Tables

**Supplementary Table 1 | Platform-level HLA typing performance comparisons.**

Summary statistics for per-sample and per-gene HLA typing accuracy across sequencing platforms. Accuracy is defined as the proportion of alleles matching the within-sample 30× HLA*LA calls at the specified resolution. Reported metrics include the mean, standard deviation (SD), and number of observations (n) per platform, providing an overview of cross-platform performance consistency.

**Supplementary Table 2 | Coriell vs GenDX concordance.**

Coriell concordance results comparing WGS-based HLA calls against GenDX reference calls. Visualized in **Figure 2D**.

**Supplementary Table 3 | SGDP platform concordance matrix.**

Pairwise concordance between platforms within the SGDP dataset. Visualized in **Supplementary Figure 1A**.

**Supplementary Table 4 | Coriell vs DRAGEN concordance matrix.**

Pairwise concordance between Coriell WGS HLA calls and DRAGEN-based calls. Visualized in **Supplementary Figure 2A**.

**Supplementary Table 5 | Cross-sample (Blood/Buccal/Saliva) platform concordance matrix.**

Pairwise concordance between platforms within the cross-sample dataset, stratified by sample source. Visualized in **Supplementary Figure 3A**.

**Supplementary Table 6 | Coriell multi-platform concordance matrix.**

Pairwise concordance between sequencing platforms within the Coriell dataset. Visualized in **Supplementary Figure 3B**.

**Supplementary Table 7 | HLA calling data of datasets analyzed in this study.**

Details for each dataset used in this work, including sequencing platform, number of samples, biological source (blood, buccal, saliva, cell line), mean genome coverage, read length, and number of HLA loci typed. This table provides context for the comparative analyses presented in Figures 3 and the Supplementary Figures.

